# Cross-neutralizing activity against Omicron could be obtained in SARS-CoV-2 convalescent patients who received two doses of mRNA vaccination

**DOI:** 10.1101/2022.02.24.22271262

**Authors:** Yukiya Kurahashi, Koichi Furukawa, Silvia Sutandhio, Lidya Handayani Tjan, Sachiyo Iwata, Shigeru Sano, Yoshiki Tohma, Hiroyuki Ohkita, Sachiko Nakamura, Mitsuhiro Nishimura, Jun Arii, Tatsunori Kiriu, Masatsugu Yamamoto, Tatsuya Nagano, Yoshihiro Nishimura, Yasuko Mori

## Abstract

The SARS-CoV-2 variant Omicron is now under investigation. We evaluated cross-neutralizing activity against Omicron in COVID-19 convalescent patients (n=23) who had received two doses of an mRNA vaccination (BNT162b2 or mRNA-1273). Surprisingly and interestingly, after the second vaccination, the subjects’ neutralizing antibody titers including that against Omicron all became seropositive, and significant fold-increases (21.1–52.0) were seen regardless of the subjects’ disease severity. Our findings thus demonstrate that at least two doses of mRNA vaccination to SARS-CoV-2 convalescent patients can induce cross-neutralizing activity against Omicron.

## BACKGROUND

The Coronavirus disease 2019 (COVID-19) pandemic caused by severe acute respiratory syndrome coronavirus 2 (SARS-CoV-2) has been a major public health problem worldwide since November 2019. As of mid-January 2022, more than 310 million individuals worldwide have been infected with SARS-CoV-2, and more than 5.5 million have died [1]. The threat posed by the SARS-CoV-2 variant Omicron (B.1.1.529) is now under intensive investigation. Omicron was first detected in Botswana on 11 November 2021 and in South Africa on 14 November 2021, and the rate of increase in the number of individuals infected with this variant has been explosive.

In the efforts to control the spread of Omicron, a major challenge has been the low seroconversion rates of cross-neutralizing antibody against Omicron in sera of the individuals who are fully COVID-19-vaccinated [2]. A booster dose of an mRNA vaccine for individuals’ acquisition of sufficient cross-neutralizing ability has been recommended by several investigators [3–5] and our research group [6], but as of this writing the data regarding the cross-neutralizing activity against Omicron in COVID-19 convalescent individuals following mRNA vaccination are lacking [5,□7,□8].

As elsewhere worldwide, the population of Japan has been attempting to overcome the COVID-19 pandemic. As of mid-January 2022, nearly 1,800,000 Japanese people have been infected with COVID-19, and approx. 18,000 people had died due to this disease [1]. The sixth wave (i.e., surge in new COVID cases) in Japan has been due to the Omicron variant. To control this pandemic in Japan, BNT162b2 vaccination (more commonly known as the Pfizer COVID-19 vaccine) was started in mid-February 2021, and mRNA-1273 vaccination (the Moderna COVID-19 vaccine) was started in mid-May 2021. As of mid-January 2022, 78.9% of Japan’s population was fully vaccinated [9].

It is important to determine whether the COVID-19 convalescent patients have cross-neutralizing activity against Omicron, because cross-neutralizing activity is the key to protection against reinfection. We thus conducted the present study to evaluate the neutralizing activity against the Delta and Omicron variants in the sera of COVID-19 convalescent patients. We also evaluated the efficacy of two doses of mRNA vaccine by analyzing the neutralizing antibody titer against Omicron in the same individuals.

## METHODS

### Diagnosis of COVID-19 and definition of severities

A positive SARS-CoV-2 antigen test or positive polymerase chain reaction (PCR) detection of the SARS-CoV-2 genome in nasal, nasopharyngeal, oropharyngeal, or saliva samples were used to confirm the diagnosis of COVID-19. We used the same definitions of severities as in our previous reports [10,□11]; ‘Asymptomatic’ patients had neither clinical symptoms nor hypoxia. ‘Mild’ patients had symptoms without evidence of pneumonia or hypoxia. ‘Moderate’ patients had clinical symptoms of pneumonia with oxygen saturation levels >90% on room air. ‘Severe’ patients suffered from pneumonia with an oxygen saturation level <90% on room air. ‘Critical’ patients needed mechanical ventilation. In the present study, we also refer to one group as ‘patients without pneumonia’; this group included the patients who did not present with pneumonia (asymptomatic and mild patients). Another group is referred to as ‘patients with pneumonia’, comprised of the patients who presented with pneumonia (moderate, severe and critical patients).

### Study site and patient recruitment

Serial sera samples of COVID-19 patients have been collected by Hyogo Prefectural Kakogawa Medical Center (Kakogawa, Hyogo, Japan) at various timepoints post-onset: 1–3, 3– 6, and 6–9 months post-onset, and approx. 12 months post-onset. In this study, the sera of patients infected between 10 March 2020 and 5 January 2021 are called ‘Term 1’, those of patients infected between 28 April 2021 and 26 May 2021 are called ‘Term 2’, and those of patients infected between 10 March 2020 and 5 January 2021 are called ‘Term 3’ excluding the patients given therapeutic monoclonal antibody drugs.

We examined the neutralizing antibody titers among SARS-CoV-2 convalescent patients. Based on the epidemiological data [12], it is likely that the patients in Term 1 were infected with D614G, those in Term 2 were infected with Alpha, and those in Term 3 were infected with Delta. We therefore compared the neutralizing antibody titers against D614G, Delta, Omicron in Term 1; we compared the titers against Alpha, Delta, Omicron in Term 2, and we compared those between Delta and Omicron in Term 3.

### Virus strains

The SARS-CoV-2 Biken-2 (B2) strain and the D614G reference variant (accession no. LC644163) received from BIKEN Innovative Vaccine Research Alliance Laboratories were used. The SARS-CoV-2 variants B.1.1.7 Alpha variant (GISAID ID: EPI_ISL_804007), B.1.167.2 Delta variant (GISAID ID: EPI_ISL_2158617), and B.1.1.529 Omicron variant (GISAID ID: EPI_ISL_7418017) were obtained from the National Institute of Infectious Diseases (NIID), Tokyo.

### Neutralization assay

The virus neutralization assay against the four SARS-CoV-2 variants D614G, Alpha, Delta, and Omicron was conducted using each authentic virus as described [10,□11] at Biosafety Level 3.

### Statistical analyses

Continuous variables are described using medians and interquartile ranges (IQRs) defined by the 25th and 75th percentiles. Categorical factors are reported as counts and percentages. The neutralizing antibody titers under two were assigned a titer of one for the geometric mean titer (GMT) calculations. The Wilcoxon signed-rank test or Friedman’s test and Bonferroni correction were performed to compare the neutralizing antibody titers. The level of statistical significance in all analyses was set at *P <* .05. Statistical analyses were performed using STATA (ver. 14.2). Sample size calculation was not performed.

### Ethics

Our study was approved by the Ethics Committees of Kobe University Graduate School of Medicine (ID: B200200) and Hyogo Prefectural Kakogawa Medical Center. Written consent or the opt-out consent for our observational study was obtained.

## RESULTS

### The patients’ characteristics

We assessed 40 sera of Term 1 patients, 12 sera of Term 2 patients, and 16 sera of Term 3 patients (**Supplementary Table 1 and 2)**. We used four groups for the sera based on the four time periods of blood sampling (1–3, 3–6, 6–9, and 12months post-onset). The numbers of patients with pneumonia were Term 1 (n=22), Term 2 (n=12), and Term 3 (n=16). The number of convalescent patients who had received two doses of a vaccine (BNT162b2 or mRNA-1273) was 19 in Term 1 and four in Term 2.

### Comparison of the neutralizing antibody titers against variants in the patients’ sera

We compared the neutralizing antibody titers against SARS-CoV-2 variants from the sera of the COVID-19 convalescent patients. These sera were collected at 1–3 months post-onset **(Figure 1A, 1B, Supplementary Figure 1, and Supplementary Table 3**). The seropositive rates of neutralizing antibody were 97.5% for D614G, 87.5% for Delta, and 37.5% for Omicron (**Figure 1A**). The GMTs of neutralizing antibody were 18.7 against D614G, 7.7 against Delta, and 1.5 against Omicron (**Supplementary Table 3**).

**Figure 1.**
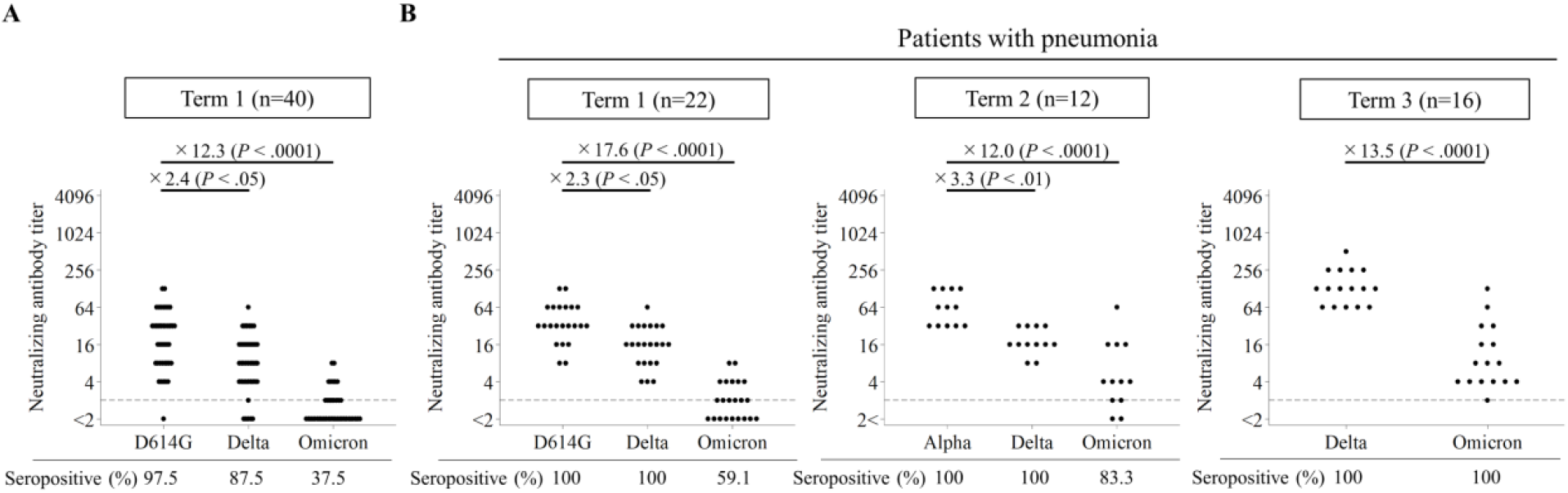
Comparison of the neutralizing antibody titers against D614G, Alpha, Delta, and Omicron from sera of convalescent COVID-19 patients. *Vertical bar:* The neutralizing antibody titer (log2 scale). *Horizontal bar:* The SARS-CoV-2 variants. All samples in panels **A** and **B** were collected at 1–3 months post-onset. **A:** The neutralizing antibody titers of the Term 1 patients. **B:** The neutralizing antibody titers in the patients with pneumonia infected during Terms 1–3. Wilcoxon signed-rank test or Friedman’s test and Bonferroni correction were performed to compare the neutralizing antibody titers. Fold-decreases of neutralizing antibody titer (Delta relative to D614G or Alpha, and Omicron relative to D614G, Alpha, or Delta) are above each graph; seropositive rates are below each graph. *Gray dash* in each graph: The cut-off titer. D614G, Alpha, and Delta are suspected to have infected the individuals in Terms 1, 2, and 3, respectively.

The results of our comparison of the neutralizing antibody titers against D614G, Delta, and Omicron in the patients with or without pneumonia in Term 1 are provided in **Supplementary Figure 1 and Supplementary Table 3**. Although the GMTs of neutralizing antibody in the patients with pneumonia in Term 1 were higher than those of the patients without pneumonia, the seropositive rate of the Term 1 patients with pneumonia was only 59.1% and the GMT of neutralizing antibody against Omicron was 2.0, the same as the cut-off point.

We next compared the neutralizing antibody titers against D614G, Alpha, or Delta with those of Omicron in the patients with pneumonia in Term 1 to Term 3 (**Figure 1B**). We aligned the severity among the patient groups because there were no sera of Term 2 or Term 3 patients without pneumonia. Significant fold-decreases of neutralizing antibody titers against Omicron relative to those against the other variants were observed: 17.6 (*P <* .0001) for Term 1, 12.0 (*P <* .0001) for Term 2, and 13.5 (*P <* .0001) for Term 3.

### Longitudinal analysis of neutralizing antibody titers against D614G, Alpha, Delta, and Omicron in patients infected in Term 1 or Term 2 after two vaccine doses

We examined the trends of neutralizing antibody titers in the patients infected during Term 1 after their two doses of vaccination at 1–3, 3–6, 6–9, and 12months post-onset (n=19) (**Figure 2A and Supplementary Table 4**). Two doses of mRNA vaccination were completed before the sera were collected at 12months post-onset. The GMTs of the neutralizing antibody against D614G and Delta tended to decline from 1–3 months post-onset to 6–9 months post-onset. The GMTs of the neutralizing antibody against Omicron were under two among three sampling points. Surprisingly and interestingly, after the two doses of vaccination, all neutralizing antibody titers including that against Omicron became seropositive and showed significant fold-increases (21.1 to 52.0) regardless of the severity of the patients’ disease (**Figures 2A and 2C, Supplementary Figure 2A, and 2B**).

**Figure 2.**
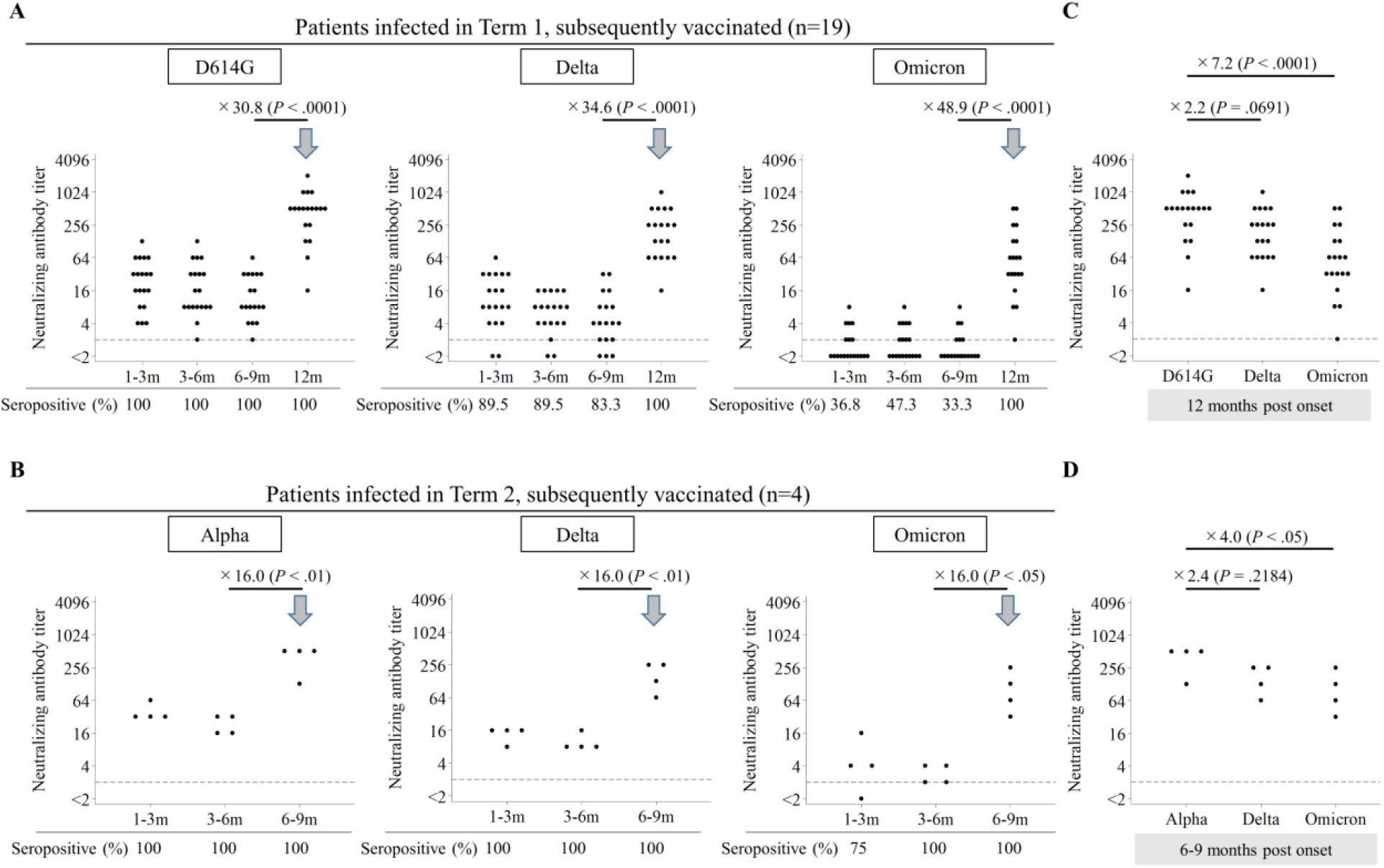
Comparison of the neutralizing antibody titers against D614G/Alpha, Delta, and Omicron in the patients infected during Term 1 or 2 and subsequently vaccinated. The neutralizing antibody titers against Delta, and Omicron in the patients infected during Term 1 and subsequently vaccinated **(A)** and the patients infected during Term 2 and subsequently vaccinated **(B). C:** Comparison of the neutralizing antibody titers against D614G, Delta, and Omicron from the same sera as in panel A collected at 12 months post-onset. **D:** Comparison of the neutralizing antibody titers against Alpha, Delta, and Omicron in the same sera as in panel **B** collected at 6–9 months post-onset. *Vertical bar:* The neutralizing antibody titer (log2 scale). *Horizontal bar:* The timing of sampling (months post-onset) (**A, B**) or the SARS-CoV-2 variants used in the virus neutralizing assay (**C, D**). *Boxes:* The SARS-CoV-2 variants. Friedman’s test and Bonferroni correction were performed to compare the neutralizing antibody titers. Fold-increases of neutralizing antibody titer (12 months post-onset relative to 6–9 months post-onset in **A**, and 6–9 months post-onset relative to 3–6 months post-onset in **B**) are shown above each graph. Fold-decreases of neutralizing antibody titer against Delta and Omicron relative to D614G are indicated in **C**, and those against Delta and Omicron relative to Alpha are indicated in **D**. Seropositive rates are shown below each graph. *Gray dash* in each graph indicates the cut-off titer. One serum at 6–9 months post-onset was missing in the group of patients infected during Term 1 and subsequently vaccinated. An *arrow* in each graph shows the timepoint of the second mRNA vaccination. D614G and Alpha are suspected to have infected the patients during Terms 1 and 2, respectively.

We then evaluated the trend of neutralizing antibody titers in the patients infected during Term 2 and subsequently vaccinated at 1–3, 3–6, and 6–9 months post-onset (n=4) (**Figures 2B, 2D, and Supplementary Table 3**). Two doses of the vaccination were completed before the sera were collected at 6–9 months post-onset. The GMTs of the neutralizing antibody against Alpha, Delta, and Omicron tended to decline from 1–3 months post-onset to 3–6 months post-onset. After two doses of the vaccination, a significant fold-increase of the neutralizing antibody titers against Omicron was also observed (**Figures 2B and 2D**), indicating that the neutralizing antibody against Omicron has been boosted by the vaccination; however, the neutralizing antibody titers against Omicron were significantly lower than those against the variants which were suspected to have infected patients (D614G or Alpha) even after vaccination (**Figure 2C, 2D, Supplementary Figure 2C, and 2D**).

## DISCUSSION

The results of our analyses revealed that the neutralizing antibody titers against Omicron in the patients infected in Term 1 were remarkably decreased (**Figure 1A, Supplementary Figure 1 and Supplementary Table 3**). Notably, the seropositive rate of neutralizing antibody titer against Omicron in the patients without pneumonia in Term 1 was only 11.1%. These data are similar to those of earlier studies [7,□8,□13]. The seropositive rate against Omicron of the patients with pneumonia was higher than that of the patients without pneumonia because a stronger immune response was elicited in the severe COVID-19 patients compared to those with mild disease (**Supplementary Figure 1**) [10].

Our findings also demonstrated significant fold-decreases against Omicron relative to D614G (×17.6), Alpha (×12.0), and Delta (×13.5) (**Figure 1B**). Another study showed clear fold-decreases against Omicron compared to Victoria (an early pandemic strain; ×16.9), Alpha (×18.4), and Delta (×25.9) [7]. In the present study, the proportion of patients whose condition was considered ‘critical’ in Term 3 (75%) was much higher than those in Term 1 (9%) and Term 2 (58%), this might have contributed to the lesser fold-decrease against Omicron relative to Delta than expected. Our results suggest that patients who have been infected with any SARS-CoV-2 variant could be possibly reinfected with Omicron.

We also observed that the neutralizing antibody titers against Omicron increased after two doses of mRNA vaccine in Term 1 and Term 2, as in other studies (**Figure 2A, 2B, Supplementary Figure 2A, 2B and Supplementary Table 4**) [5,□13]. It was reported that the neutralizing antibody activity against Omicron in convalescent patients who were subsequently vaccinated was around ten times higher than that in fully vaccinated patients without past infection [5,□13]. In the present investigation, the ratios of the neutralizing antibody titer against Omicron relative to the other variants (D614G or Alpha) from vaccinated patients after their recovery (**Figure 2C and 2D**) were almost the same as those against Omicron compared to the D614G in uninfected individuals who received three doses of COVID-19 mRNA vaccine [4,□8,□13]. Although it remains unknown whether one dose is sufficient for convalescent patients with severe symptoms and whether a third dose is necessary for convalescent patients with mild symptoms, our results show that at least two doses of mRNA vaccination to SARS-CoV-2 convalescent patients may induce cross-neutralizing activity against Omicron comparably to three doses of mRNA vaccination to uninfected individuals.

## Data Availability

All data produced in the present work are contained in the manuscript.

## Footnotes

### Conflict of interest

The authors declare no conflicts of interest associated with this manuscript.

## Funding/Support

This work was supported by Hyogo Prefectural Government and the Kansai Economic Federation (KANKEIREN). The funders had no role in this study.

## Meeting

This study has never been presented anywhere.

## Acknowledgments

We thank Kazuro Sugimura MD, PhD (Superintendent, Hyogo Prefectural Hospital Agency and Professor, Kobe University) for his full support to promote this study. We express our sincere gratitude for cooperation and participation of staffs of Hyogo Prefectural Kakogawa Medical Center. We thank for BIKEN Innovative Vaccine Research Alliance Laboratories providing SARS-CoV-2 B2 strain. We thank the National Institute of Infectious Disease Japan for providing SARS-CoV-2 Alpha, Delta, and Omicron variants. YK obtained Taniguchi Memorial Scholarship program founded by BIKEN Foundation. SS^1^ and LT got Japanese Government (Monbukagakusho:MEXT) Scholarships.

## Author Contributions

All authors contributed to the concept of this article. YK drafted the manuscript; YM provided revisions; YK, JA, MN, and YM analyzed the data; YK, KF, SS^1^, and LT did the experiments; YM and YN supervised the experiments; SI, YT, SS^3^, SN, TK, MY and TN collected the samples; YM conducted the project; All authors approved the final version of the manuscript

---

**Supplementary Figure 1.**
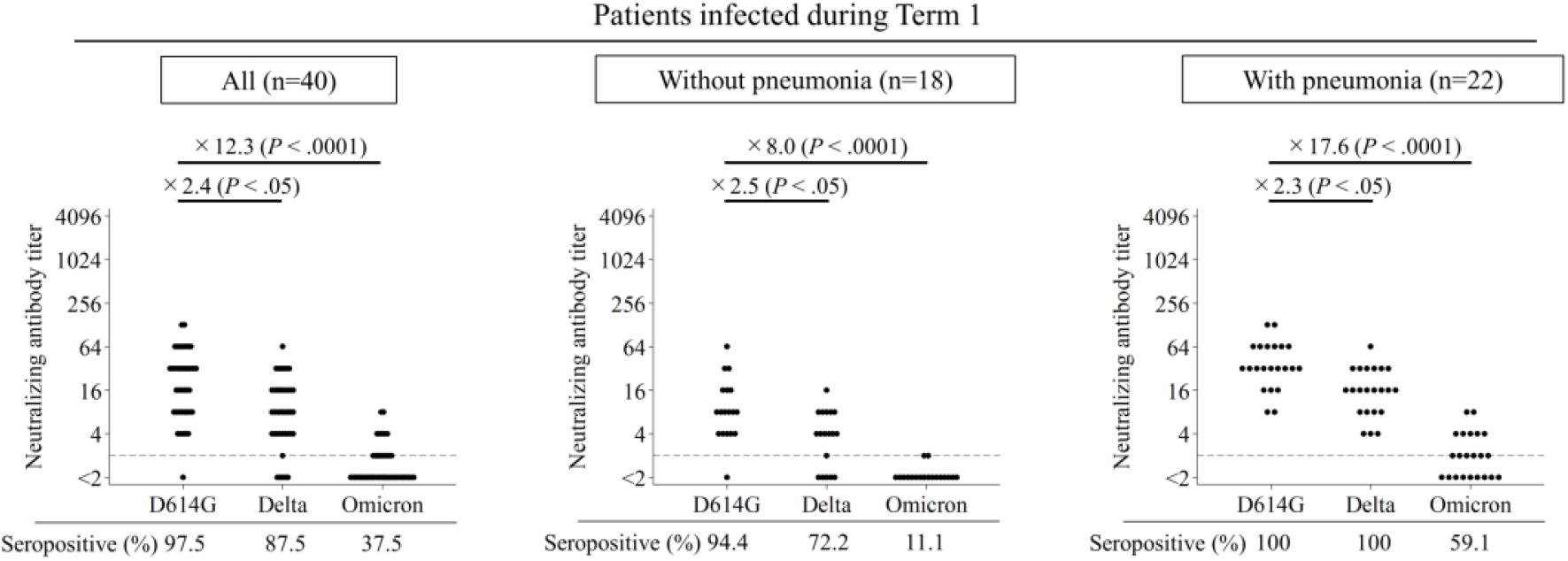
Comparison of the neutralizing antibody titers against D614G, Delta, and Omicron in the patients infected during Term 1, by severity. *Vertical bar:* The neutralizing antibody titer (log2 scale). *Horizontal bar:* The SARS-CoV-2 variants. All samples were collected at 1–3 months post-onset. The neutralizing antibody titer in the patients infected during Term 1, those without pneumonia in Term 1, and those with pneumonia in Term 1 are shown. Friedman’s test and Bonferroni correction were used to compare the neutralizing antibody titers. Fold-decreases of neutralizing antibody titer (Delta relative to D614G, and Omicron relative to D614G) are shown above each graph. Seropositive rates are shown below each graph. *Gray dash* in each graph indicates the cut-off titer. D614G is thought to have infected the patients during Term 1.

**Supplementary Figure 2.**
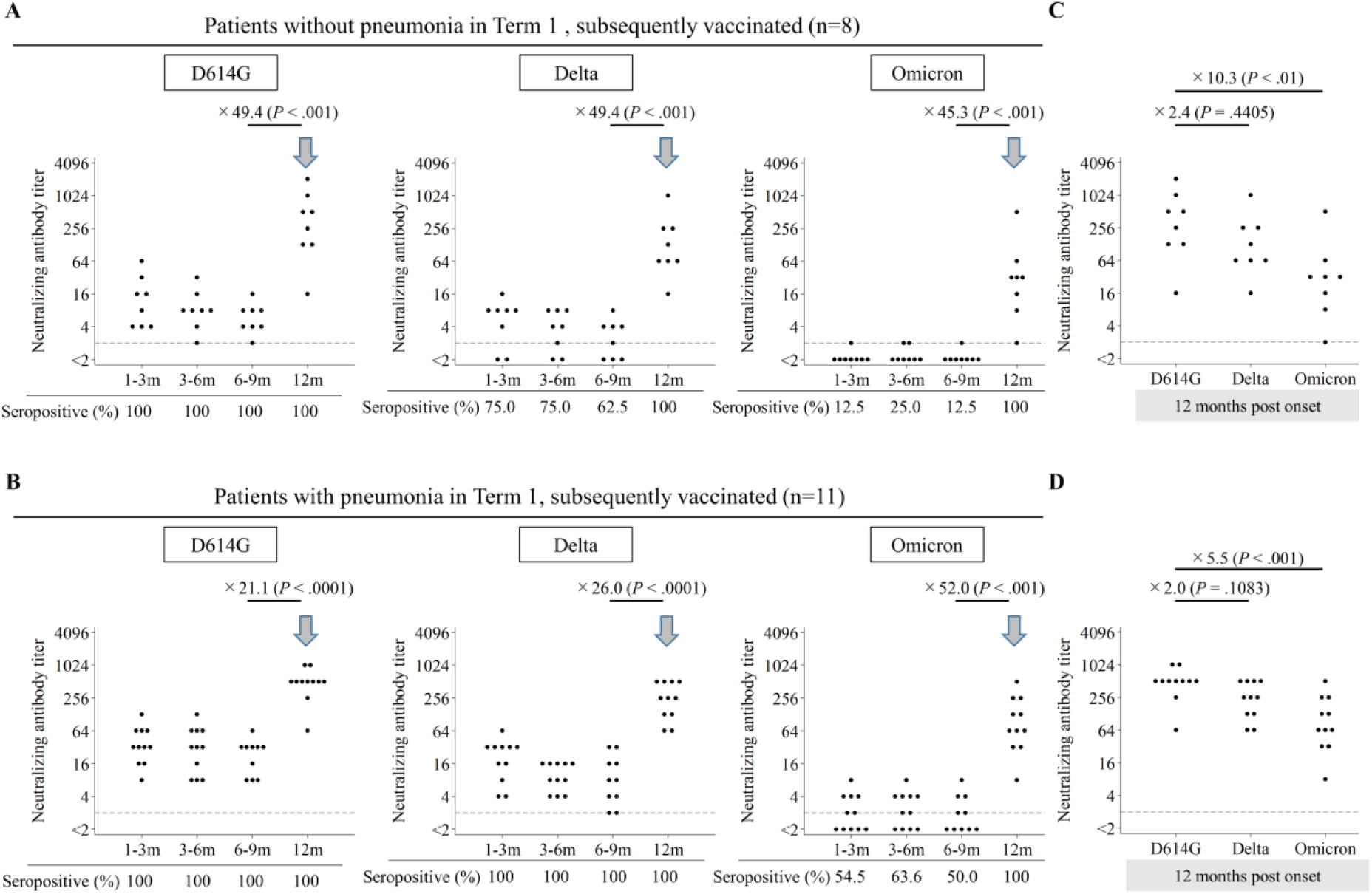
Comparison of the neutralizing antibody titers against D614G, Delta, and Omicron in the patients infected during Term 1 and subsequently vaccinated, by severity. Comparison of the neutralizing antibody titers against D614G, Delta, and Omicron in the patients without pneumonia in Term 1 and subsequently vaccinated **(A)** and in the patients with pneumonia in Term 1 and subsequently vaccinated **(B)**. Comparison of the neutralizing antibody titers against D614G, Delta, and Omicron in the same sera as in panel A collected at 12 months post-onset **(C)**. Comparison of the neutralizing antibody titers against Alpha, Delta, and Omicron in the same sera as in panel **B** collected at 12 months post-onset **(D)**. *Vertical bar:* The neutralizing antibody titers (log2 scale). *Horizontal bar:* The timing of sampling (months post-onset) (**A, B**) or SARS-CoV-2 variants used in the virus neutralizing assay (**C, D**). *Boxes:* The SARS-CoV-2 variants. Friedman’s test and Bonferroni correction were performed to compare the neutralizing antibody titers. Fold-increases of the neutralizing antibody titers (12 months post-onset relative to 6–9 months post-onset in **A**, and 6–9 months post-onset relative to 3–6 months post-onset in **B**) are shown above each graph. Fold-decreases of the neutralizing antibody titers against Delta and Omicron relative to D614G are indicated in **C** and **D**. Seropositive rates are shown below each graph. *Gray dash* in each graph indicates the cut-off titer. One serum at 6–9 months post-onset was missing in the group of patients infected during Term 1 and subsequently vaccinated. An *arrow* in each graph indicates the timepoint of the second mRNA vaccination. D614G is thought to have infected the patients during Term 1.

**Supplementary Table 1.**
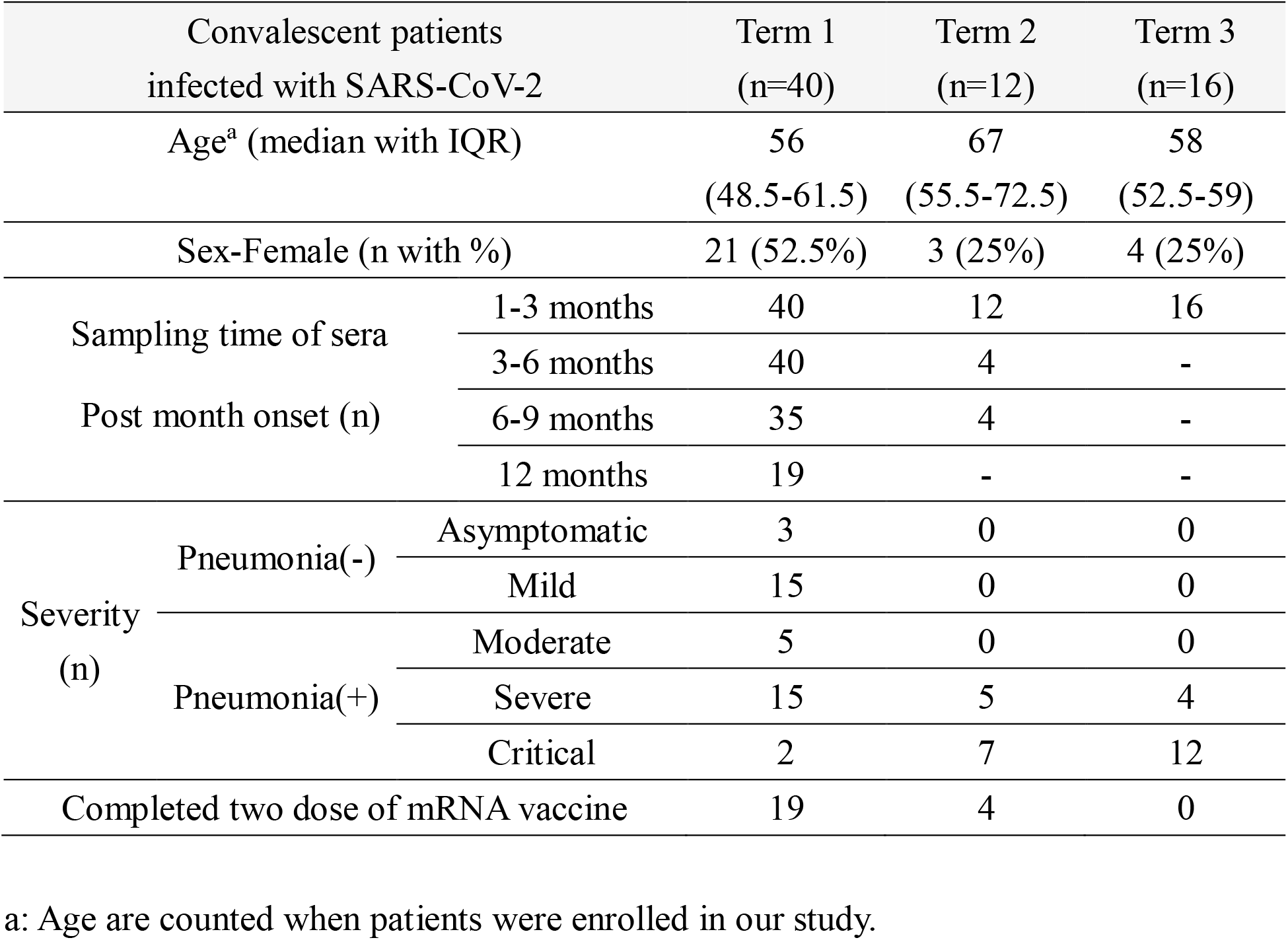
Characteristics of convalescent patients.

**Supplementary Table 2.** Median days post onset of sampling sera for SARS-CoV-2 convalescent patients.

| Sera | Sampling time (month post onset) | Median days with IQR |  |
| --- | --- | --- | --- |
| Term 1 | 1-3m (n=40) | 46.5 | (42.5-54) |
|  | 3-6m (n=40) | 116.5 | (109-132) |
|  | 6-9m (n=35) | 202 | (197-215) |
|  | 12m (n=19) | 379 | (368-395) |
| Term 2 | 1-3m (n=12) | 41 | ( 34.5-48) |
|  | 3-6m (n=4) | 108.5 | (100.5-119) |
|  | 6-9m (n=4) | 199.5 | (192.5-209) |
| Term 3 | 1-3m (n=21) | 43 | (39-51) |

**Supplementary Table 3.** Geometric mean titers (GMTs) of neutralizing antibody in sera of SARS-CoV-2 convalescent patients at 1-3 months onset.

| Sera | Variant | GMT of neutralizing antibody<sup>a</sup> |  |
| --- | --- | --- | --- |
| Term 1<br>(n=40) | D614G | 18.7 | (13.1-26.6) |
|  | Delta | 7.7 | ( 5.4 -11.0) |
|  | Omicron | 1.5 | ( 1.2 - 1.9) |
| Term 1<br>without pneumonia (n=18) | D614G | 8.6 | ( 5.3 -14.0) |
|  | Delta | 3.4 | ( 2.2 - 5.4) |
|  | Omicron | 1.1 | ( 1.0 -1.2) |
| Term 1<br>with pneumonia (n=22) | D614G | 35.2 | (25.2-49.0) |
|  | Delta | 15.0 | (10.7-21.1) |
|  | Omicron | 2.0 | ( 1.5 -2.7) |
| Term 2<br>with pneumonia (n=12) | Alpha | 60.4 | (40.6-89.8) |
|  | Delta | 18.0 | (13.1-24.6) |
|  | Omicron | 5.0 | ( 2.3-11.3) |
| Term 3<br>with pneumonia (n=16) | Delta | 133.7 | (94.9-188.4) |
|  | Omicron | 9.9 | ( 5.3 -18.6) |

a: Neutralizing antibody titers under two were assigned a titer of one for geometric mean titer (GMT) calculations. GMTs were shown with 95% CI.

**Supplementary Table 4.**
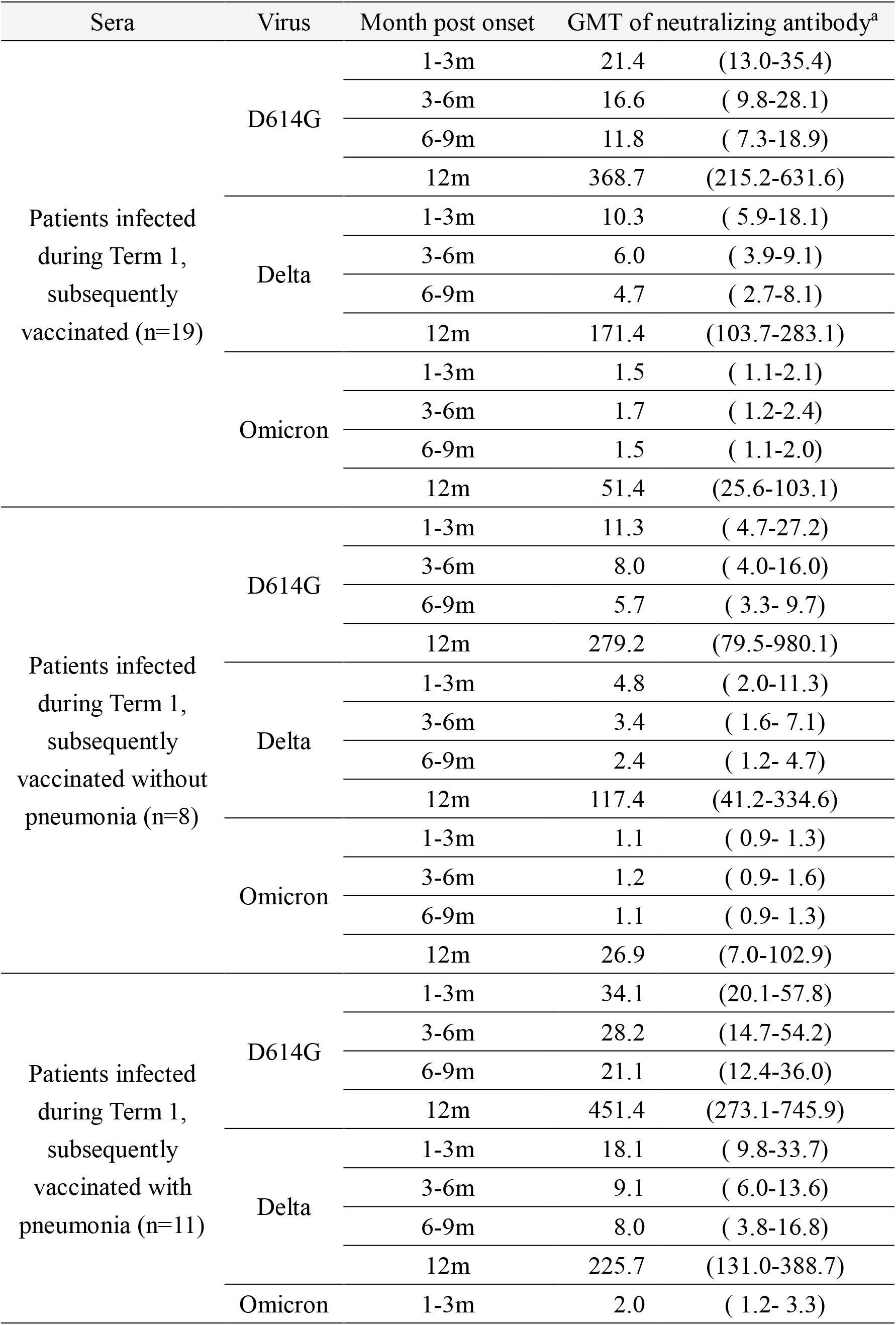
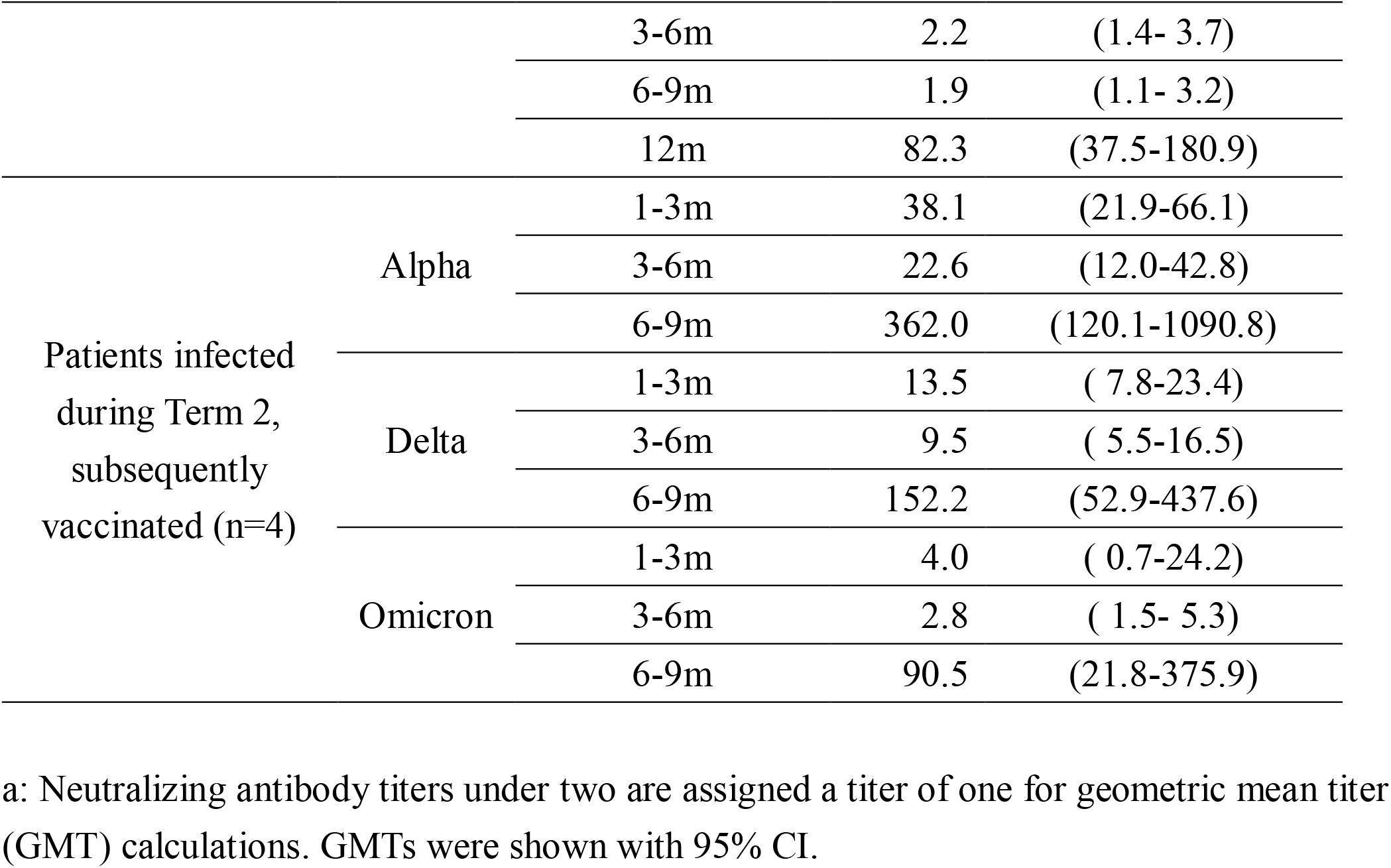
Geometric mean titers (GMTs) of neutralizing antibody in sera of SARS-CoV-2 convalescent, subsequently vaccinated patients.

| Sera | Virus | Month post onset | GMT of neutralizing antibody<sup>a</sup> |  |
| --- | --- | --- | --- | --- |
| Patients infected during Term 1, subsequently vaccinated (n=19) | D614G | 1-3m | 21.4 | (13.0-35.4) |
|  |  | 3-6m | 16.6 | ( 9.8-28.1) |
|  |  | 6-9m | 11.8 | ( 7.3-18.9) |
|  |  | 12m | 368.7 | (215.2-631.6) |
|  | Delta | 1-3m | 10.3 | ( 5.9-18.1) |
|  |  | 3-6m | 6.0 | ( 3.9-9.1) |
|  |  | 6-9m | 4.7 | ( 2.7-8.1) |
|  |  | 12m | 171.4 | (103.7-283.1) |
|  | Omicron | 1-3m | 1.5 | ( 1.1-2.1) |
|  |  | 3-6m | 1.7 | ( 1.2-2.4) |
|  |  | 6-9m | 1.5 | ( 1.1-2.0) |
|  |  | 12m | 51.4 | (25.6-103.1) |
| Patients infected during Term 1, subsequently vaccinated without pneumonia (n=8) | D614G | 1-3m | 11.3 | ( 4.7-27.2) |
|  |  | 3-6m | 8.0 | ( 4.0-16.0) |
|  |  | 6-9m | 5.7 | ( 3.3- 9.7) |
|  |  | 12m | 279.2 | (79.5-980.1) |
|  | Delta | 1-3m | 4.8 | ( 2.0-11.3) |
|  |  | 3-6m | 3.4 | ( 1.6- 7.1) |
|  |  | 6-9m | 2.4 | ( 1.2- 4.7) |
|  |  | 12m | 117.4 | (41.2-334.6) |
|  | Omicron | 1-3m | 1.1 | ( 0.9- 1.3) |
|  |  | 3-6m | 1.2 | ( 0.9- 1.6) |
|  |  | 6-9m | 1.1 | ( 0.9- 1.3) |
|  |  | 12m | 26.9 | (7.0-102.9) |
| Patients infected during Term 1, subsequently vaccinated with pneumonia (n=11) | D614G | 1-3m | 34.1 | (20.1-57.8) |
|  |  | 3-6m | 28.2 | (14.7-54.2) |
|  |  | 6-9m | 21.1 | (12.4-36.0) |
|  |  | 12m | 451.4 | (273.1-745.9) |
|  | Delta | 1-3m | 18.1 | ( 9.8-33.7) |
|  |  | 3-6m | 9.1 | ( 6.0-13.6) |
|  |  | 6-9m | 8.0 | ( 3.8-16.8) |
|  |  | 12m | 225.7 | (131.0-388.7) |
|  | Omicron | 1-3m | 2.0 | ( 1.2- 3.3) |
|  |  | 3-6m | 2.2 | (1.4- 3.7) |
|  |  | 6-9m | 1.9 | (1.1- 3.2) |
|  |  | 12m | 82.3 | (37.5-180.9) |
| Patients infected during Term 2, subsequently vaccinated (n=4) | Alpha | 1-3m | 38.1 | (21.9-66.1) |
|  |  | 3-6m | 22.6 | (12.0-42.8) |
|  |  | 6-9m | 362.0 | (120.1-1090.8) |
|  | Delta | 1-3m | 13.5 | ( 7.8-23.4) |
|  |  | 3-6m | 9.5 | ( 5.5-16.5) |
|  |  | 6-9m | 152.2 | (52.9-437.6) |
|  | Omicron | 1-3m | 4.0 | ( 0.7-24.2) |
|  |  | 3-6m | 2.8 | ( 1.5- 5.3) |
|  |  | 6-9m | 90.5 | (21.8-375.9) |

a: Neutralizing antibody titers under two are assigned a titer of one for geometric mean titer (GMT) calculations. GMTs were shown with 95% CI.

## Notes

### Competing Interest Statement

The authors have declared no competing interest.

### Funding Statement

This study was funded by Hyogo prefectural government.

### Author Declarations

Ethics committee of Kobe University gave ethical approval for this work.

